# A Prediction Model of PTSD in the Israeli Population in the Aftermath of October 7^th^, 2023, Terrorist Attack and the Israel-Hamas War

**DOI:** 10.1101/2024.02.25.24303235

**Authors:** Dana Katsoty, Michal Greidinger, Yuval Neria, Aviv Segev, Ido Lurie

**Affiliations:** Psychology Department, The Hebrew University of Jerusalem, Jerusalem, Israel; Department of Counseling and Human Development, Faculty of Education, University of Haifa, Haifa, Israel; Department of Psychiatry, Columbia University Irving Medical Center, New York, New York, USA; New York State Psychiatric Institute, Columbia University Irving Medical Center, New York, USA; Shalvata Mental Health Center, Hod Hasharon, Israel; School of Medicine, Faculty of Medical and Health Sciences, Tel Aviv University, Tel Aviv, Israel; Department of Psychosis Studies, Institute of Psychiatry, Psychology and Neuroscience, Kings College London, London, UK

**Keywords:** post-traumatic stress disorder (PTSD), war, terror, trauma, epidemiology, prediction model, mental health services

## Abstract

**Background:** On October 7^th^, 2023, a mass terror attack was launched by Hamas militants, which was followed by the Israel-Hamas war. These events constitute a nationwide trauma with major ramifications for public mental health. This article presents an evidence-based model for the prediction of the prevalence of posttraumatic stress disorder (PTSD) related to the terrorist attack and the war.

**Main body:** The creation of the model consisted of several steps. Firstly, the Israeli population was divided into six groups based on the intensity, context, and type of traumatic exposure (direct exposure to terror, close proximity to terror, soldiers in combat and support units, intense exposure to rocket attacks, moderate exposure to rocket attacks, and indirectly affected communities), and the population size of each group was assessed using official national databases. Secondly, an estimation of the expected prevalence of PTSD in each of the exposure groups was based on a review of the relevant literature. A random-effects meta-analysis of the prevalence rates was conducted separately per each exposure group. Finally, the predicted number of PTSD causalities in the population was calculated by multiplying the group size and the PTSD prevalence estimation. Based on population size and estimated PTSD prevalence within each exposure category, the model predicts that approximately 5.3% (N=519,923) of the Israeli population (95% prediction interval, 160,346-879,502), may develop PTSD as a result of the terrorist attack and the war.

**Conclusions:** The predicted number of individuals with PTSD following mass trauma is expected to be considerable. The presented model can assist policymakers, clinicians, and researchers in preparing and devising adequate interventions for the mental health needs of large populations. Moreover, this model can be applied in other instances of mass-trauma exposure.

## BACKGROUND

The terrorist attack conducted by Hamas on October 7^th^, 2023, has resulted in nationwide mass trauma, with major implications for public mental health across large groups within the population, as it ranks one of the deadliest terrorist attacks since 1970 in absolute number of fatalities (Statista, 2023). On that day, Hamas militants infiltrated Israel and targeted communities and army bases surrounding the Gaza Strip. This assault resulted in a mass tragedy, claiming the lives of over 1,100 Israeli civilians, soldiers, and foreign citizens, with 248 people abducted and taken to the Gaza Strip, including infants, children, women, and elderly people. Thousands of civilians were injured, and many were subject to torture and sexual violence. Within the first 4 hours of the attack, 3,000 rockets were fired into Israel. The attack initiated an ongoing war between Israel and Hamas that had devastating effects on civilians both in Israel and in Gaza, including mass casualties, injuries, and displacement of civilians (Devi, 2023; Fox and Kolitz, 2023; Gavrielov, 2023; Peled and Jones, 2023; Statista, 2023; Saidel et al., 2024). Many of the atrocities were also filmed and virally spread online through social media (Cortellessa, 2023; Impelli, 2023).

It is crucial to assess the expected psychiatric outcomes of such atrocities, as exposure to war and terror has detrimental implications for the mental health of both individuals and communities (e.g., Neria et al. 2010; Neria et al., 2011; Hoppen, et al., 2021). One of the most prominent outcomes of such exposure is post-traumatic stress disorder (PTSD; (5th ed., DSM-5; American Psychiatric Association, 2013)). The burden of PTSD is substantial, with high direct and indirect healthcare costs (Davis, et al., 2022; von der Warth, et al., 2020), and even more so when the disorder is untreated (e.g., Pityaratstian et al., 2015). A plethora of research assessed the prevalence of PTSD as a result of exposure to war and terrorism and has demonstrated a notable dose-response relationship between the type and intensity of exposure and the risk for PTSD (e.g., Johnson & Thompson, 2008; May & Wisco, 2016; Neria, et al., 1998; Neria, et al., 2006; North, 2011). Yet, while most terrorist attacks involve a segment of the population, in the October 7^th^ attack and the Israel-Hamas war the entire Israeli population was exposed to the atrocities to some extent.

To systematically estimate the risk for PTSD across Israeli society, we developed an evidence-based predictive model for the prevalence of PTSD across different groups of the population, depending on the varying levels of exposure to the October 7^th^ terror attacks and the Israel-Hamas war. This prediction model for PTSD prevalence in populations exposed to large-scale trauma could be applied in future occasions of mass trauma events, with relevant adjustments in terms of exposure type and extent.

## MAIN TEXT

### Methods

#### Procedure

The development of the prediction model consisted of the following steps: 1) Classifying exposure groups across the Israeli population, based on different elements of traumatic exposure, i.e.,, the context in which exposure occurred, the intensity of exposure to the attacks, and the type of exposure – direct exposure (e.g., a terrorist attack), or other indirect forms of exposure (e.g., rocket attacks). The size of each of these exposure groups was estimated according to official government information, media publications and national open-access information (see Appendix 1); 2) Estimating the expected prevalence rates of PTSD in each of the exposure groups. This was done in three steps: Firstly, we conducted a literature review concerning the prevalence of PTSD following exposure to war and terror, by searching for reviews, systematic reviews, and meta-analyses in PubMed using the terms: (PTSD OR post-traumatic stress disorder) AND (prevalence OR incidence OR rate*) and (war OR terror), as well as relying on Google Scholar for an additional search. For each exposure group, we attempted to identify review papers that reflect the nature of the traumatic exposure of the specific group. Secondly, we examined all studies within each review and omitted studies that did not meet the inclusion criteria (that are addressed under the methodological challenges section). Thirdly, we conducted a random-effects meta-analysis of the prevalence rates separately per each group, based on all studies that met the inclusion criteria within each review. The meta-analysis aggregated the proportion of PTSD cases out of each sample, across all studies (i.e., comparison between groups with respect to a dichotomous dependent variable, which is PTSD prevalence); 3) Calculating the predicted number of PTSD causalities in the population by multiplying the group size and the estimated prevalence of PTSD.

#### Classifying Exposure Groups

1. **Direct exposure to the October 7^th^ terrorist attacks.** These attacks included two main areas: the music festivals in Kibbutz Reim, and communities and cities around the Gaza Strip.
2. **Close proximity to the October 7^th^ terrorist attacks**. Communities and areas within cities in this group were not infiltrated by Hamas perpetrators, but their residents perceived that their lives were severely threatened due to the invasion of nearby areas, and the high risk of the perpetrators invading their homes. Many of these civilians were hiding in shelters for hours or even days during and after the attack (Awad & Lubell, 2023).
3. **Soldiers in combat and support units involved in the war.** This group includes soldiers on active duty and reservists. Both infantry units and soldiers in support units, which were not directly involved in combat, were included in this group.
4. **Civilians under intense exposure to rocket attacks (living up to 40 kilometers (25 miles) from the Gaza Strip).** Civilians who live in the cities and communities located up to 40 km from the Gaza-Israel border and have been under recurring rocket attacks. The delineation of 40 km, as utilized in various studies (Besser et al., 2013; Besser et al., 2014; Besser et al., 2015; Gil et al., 2015; Gil et al., 2016; Weinberg et al., 2015), was selected based on the distinct characteristics of this area, which involves a brief window of time to reach shelters immediately after the rocket siren is activated (up to 1 minute; Israel Home Front Command, 2023). This group was also exposed to the largest number of rocket attacks (Israel Home Front Command, 2023).
5. **Civilians under moderate exposure to rocket attacks (living 40-80 km (25-50 miles) from the Gaza Strip).** Civilians living between 40-80 kilometers from the Israel-Gaza border who have been exposed to rocket attacks to a substantial degree, but to a smaller extent compared to the previous group, and with a longer duration of time to enter shelters (1 to 1.5 minutes; Israel Home Front Command, 2023).
6. **Indirectly affected communities** (**living more than 80 km (50 miles) from the Gaza Strip).** Civilians who live in communities and cities located more than 80 km from the Israel-Gaza border. Some of them have been exposed to rocket attacks, but to a substantially smaller degree, which we considered less threatening and interfering with everyday life. Additionally, they have been exposed to the traumatic events through media coverage. To note, this group includes the rest of the Israeli population that was not included in the previous categories.

#### Addressing Methodological challenges

The development of the predictive model required addressing several significant methodological and theoretical challenges, each demanding a distinctive solution:

### Variability in PTSD prevalence estimates

Past research has demonstrated striking differences across studies assessing PTSD prevalence. This variability was attributed to methodological differences among the studies, such as differences in the method of PTSD measurement (i.e., clinical assessments vs. self-report questionnaires), time of measurement after the trauma, and specific characteristics of the exposure (e.g., Sundin et al., 2010; Hoffman et al., 2011).

### Method of PTSD measurement

Two methods of measurement are used to determine the occurrence of PTSD. The first is clinical assessments, which include validated diagnostic interviews with explicit PTSD diagnostic criteria based on the Diagnostic and Statistical Manual of Mental Disorders (5^th^ ed., text rev.; DSM-5-TR; American Psychiatric Association) or the International Disease Classification (11^th^ ed.; ICD-11; World Health Organization, 2019). The second method is self-report questionnaires or symptom checklists, wherein individuals are classified within a category of probable PTSD when exceeding a specific questionnaire cutoff (e.g., Neria, et al., 2011). These two methods of measurement have been shown to yield significantly different PTSD prevalence estimates, and self-report questionnaires have been argued to overestimate the prevalence of PTSD (Turner et al., 2003). More generally, as self-report questionnaires or symptom checklists do not properly assess the severity, frequency, duration, covariation, and degree of interference (McNally, et al., 2003), and since clinical assessments are generally considered the gold standard for PTSD diagnosis, we preferred clinical assessments in the literature search process. In case a review included both clinical assessments and questionnaires, we omitted the questionnaire estimates and included only clinical assessments in the meta-analysis. In instances where no studies using clinical assessment were included in a review, we implemented a mitigating correction factor in order to maintain the most conservative estimates. A factor of 0.71 was chosen as self-rating and symptom checklists were found to overestimate the prevalence of PTSD by 41% (Steel, et al., 2009). Importantly, in questionnaire measurements, we omitted studies that did not assess PTSD specifically in relation to war/terror exposure or did not explicitly state that the measurement was conducted with regard to this specific form of exposure.

### Time of measurement after trauma exposure

Although PTSD can only be diagnosed one month or more after the traumatic exposure, several studies have examined PTSD symptoms within the time frame of the traumatic exposure, or less than one month after the exposure. These assessments reflect acute stress disorder (ASD) symptomatology and not PTSD (American Psychiatric Association, 2013). Therefore, studies that examined post-traumatic symptoms less than one month after traumatic exposure were omitted from analyses. Additionally, in case a specific study included more than one measurement, we included only the highest prevalence estimate (as long as that measurement was conducted one month or more after the traumatic exposure). Importantly, we only included current, point-prevalence estimates, as opposed to period prevalence, to avoid inflation of estimates.

### The unique nature of exposure and implications for the literature review process

The events of the war were characterized by unique types of exposure to terror and violence. Therefore, for some of the exposure groups, we were not able to find a review that reflected the nature of exposure. This was the case for two groups: group 2 (close proximity to the October 7^th^ terror attacks), and group 5 (moderate exposure to rocket attacks). To derive estimates for these two groups, we relied on estimates from the other exposure groups, and addressed the specific manner in which the estimate was calculated.

### Prevalence rates in children vs. adults

Children may respond to trauma differently compared to adults (Shaw, 2003). The majority of studies conducted regarding the prevalence of PTSD focused on adults, and studies that compared PTSD prevalence in adults and children demonstrated conflicting results (Tang, et al., 2017, Fairbank & Fairbank, 2009, Pat-Horenczyk, et al., 2015). Due to the ambiguity of these estimations, we did not distinguish between children and adults within each exposure group.

### Other issues

Notably, many individuals in each group had more than one form of exposure (e.g., survivors of the October 7^th^ attack were also subject to rocket attacks). In cases of multifaceted exposure, we chose the highest prevalence rate. Additionally, we omitted studies that examined only populations seeking mental health services and studies conducted in clinical settings, as these are expected to include higher PTSD prevalence compared to general samples of people who were exposed to trauma.

### Statistical analysis

Meta-analyses were conducted using R package metafor (Viechtbauer, 2010).

## Results

We present the identified reviews and address the number of studies that were omitted within each review (if such omission was conducted). The supplementary material includes the full list of excluded studies within each review. We then report the results of the meta-analyses for groups in which estimates were derived from meta-analyses (including prevalence estimates, confidence intervals, which are addressed as prediction intervals, and heterogeneity estimates). The supplementary material includes forest plots of summary effects for each of the meta-analyses. We also describe how group sizes were determined (with a full elaboration in Appendix 1) and present the number and range of people within each group who are expected to develop PTSD, calculated by multiplying the estimate and confidence interval by the group size. For groups in which estimates were not derived from meta-analyses, we address how the estimate was determined and address the determination of group size, estimate, and range.

Table 1 summarizes the results of the prediction model separately for each exposure group described above – the population size estimate, the probability of developing PTSD (prevalence estimate and range), and the expected number of people with PTSD (the number of people within each group multiplied by the prevalence estimate).

**Table 1:** Results of the prediction model.

| **Exposure group** | **Group Size**<br>**(after**<br>**exclusions)** | **PTSD**<br>**prevalence**<br>**range** | **PTSD**<br>**prevalence**<br>**estimate** | **Expected**<br>**N- PTSD** | **Lower**<br>**bound N** | **Upper**<br>**bound N** | **Reference** | **Data Type** |
| --- | --- | --- | --- | --- | --- | --- | --- | --- |
| **1) Direct exposure to the**<br>**October 7th terrorist attacks** | 39,664 | 0.21-0.42 | 0.32 | 12,564 | 8,474 | 16,654 | Paz García-Vera et<br>al., 2016. | Clinical<br>assessments |
| **2) Close proximity to the**<br>**October 7th terrorist attacks** | 121,061 | 0.03-0.17 | 0.10 | 12,354 | 3,744 | 20,964 | - | Modeling |
| **3) Soldiers in combat and**<br>**support units involved in the**<br>**war** | 144,227 | 0.01-0.14 | 0.08 | 11,021 | 1,494 | 20,549 | Richardson et al.,<br>2010; Xue, et al.,<br>2015 | Clinical<br>assessments |
| **4) Civilians under intense exposure to rocket attacks (living up to 40 kilometers (25 miles) from the Gaza Strip)** | 1,069,011 | 0.03-0.17 | 0.10 | 109,088 | 33,058 | 185,118 | Greene et al., 2018 | Adjusted Probable PTSD |
| **5) Civilians under moderate exposure to rocket attacks (living 40-80 km (25-50 miles) from the Gaza Strip)** | 4,960,469 | 0.02 - 0.10 | 0.06 | 304,182 | 92,166 | 516,198 | - | Modeling |
| **6) Indirectly affected communities (living more than 80 km (50 miles) from the Gaza Strip)** | 3,433,286 | 0.01-0.03 | 0.02 | 70,714 | 21,410 | 120,019 | Paz García-Vera et al., 2016; Greene et al., 2018 | Clinical assessments |
| **Total** |  |  |  | 519,923 | 160,346 | 879,502 |  |  |

### Group 1: Direct exposure to the October 7^th^ terror attacks

We relied on a systematic review by Paz García-Vera, et al. (2016), which examined several types of exposure to terrorism, one of which was direct exposure to terror attacks. All studies presented in this review included clinical assessments.

#### Omission of studies

One study was omitted as it included a help-seeking population. As mentioned earlier, in longitudinal studies we included measurements with the highest prevalence.

#### Meta-analysis of included studies

Nine studies were included in the meta-analysis. The mean prevalence estimate was μ^ = 0.32 (95%CI 0.21-0.42). The Q-test indicated that outcomes are heterogeneous, suggesting that different studies vary in the observed prevalence estimate. Moreover, the I2 statistic indicated that a meaningful portion of the variance could be explained by true heterogeneity rather than chance (Q(9) = 233.79, p < 0.001, I2 = 93.86%, τ = .15).

#### Group size and numerical estimate and range

We included communities and areas within cities that were infiltrated by Hamas perpetrators, a total of 39,664 people (See Appendix 1). After multiplying the estimate and CI by group size, we predict that 12,564 (a range of 8,474 to 16,654) people are expected to develop PTSD.

### Group 3: Soldiers in combat and support units involved in the war

We identified two literature reviews that were relevant for this exposure group. Both reviews examined combat-PTSD after war exposure (Richardson, et al, 2010; Xue, et al., 2015). These reviews included both clinical assessments and questionnaire assessments.

#### Omission of studies

In the review by Richardson, et al. (2010) we omitted studies for the following reasons: 18 studies included questionnaire assessments or non-clinical assessments; two studies re-analyzed existing data from another sample; one study included help-seeking clinical population; and one study that did not examine current PTSD prevalence. In the review by Xue, et al., (2015), we omitted studies for the following reasons: 23 studies included questionnaire assessments or non-clinical assessments; three studies examined PTSD diagnosis in medical files without specific reference to the war exposure; and three studies did not examine current PTSD prevalence.

#### Meta-analysis of included studies

Four studies were included in the meta-analysis. The mean prevalence estimate was μ^ = 0.08 (95%CI 0.01-0.14). The Q-test and I2 statistic indicated that outcomes are heterogeneous and that a meaningful portion of the variance could be explained by true heterogeneity rather than chance (Q(3) = 226.18, p < 0.001, I2 = 99.38%, τ = .06).

#### Group size and numerical estimate and range

An estimate for the group size was made based on reports in the media of 10 divisions operating in the escalation areas: the Gaza Strip, the Israel-Lebanon border, and the West Bank (See Appendix 1). The size of a division ranges from 13,000 to 20,000 soldiers, with an average of 16,500, and a total of 165,000 people (Hamichlol, 2023). Importantly, the 20,773 soldiers that were subtracted from the soldiers’ group were classified under groups with higher prevalence rates, since in cases of multifaceted exposure, individuals were classified under the higher-prevalence group (see Appendix 1 for an elaboration). After this subtraction, the group size was 144,227 people. After multiplying the estimate and CI by the group size, we predict that 11,021 (a range of 1,494 to 20,549) people are expected to develop PTSD.

### Group 4: Civilians under intense exposure to rocket attacks (living up to 40 km from the Gaza Strip)

We relied on a review by Greene et al (2018), which examined PTSD rates in residents of the south of Israel that were under exposure to continuous rocket attacks. This review only included questionnaire-based assessments, and therefore all estimates included in the review were multiplied by the correction factor (of 0.71).

#### Omission of studies

Twelve of the studies presented in the review included PTSD prevalence estimates. Out of these studies, we omitted studies for the following reasons: two studies examined PTSD within the timeframe of war exposure; one study did not specify whether measurement was conducted during or after war exposure; and four studies did not clarify whether they assessed symptoms that related specifically to rocket attacks.

#### Meta-analysis of included studies

Five studies (and six samples) were included in the meta-analysis. The mean prevalence estimate was μ^ = 0.10 (95% CI[0.03-0.17]). The Q-test and I2 statistic indicated that outcomes are heterogeneous and that a meaningful portion of the variance could be explained by true heterogeneity rather than chance (Q(5) = 116.43 p < 0.001, I2 = 97.46%, τ = .09).

#### Group size and numerical estimate and range

This group amounted to a total of 1,069,011 people (See Appendix 1). Groups 1 and 2, which are also located within 40 km from Gaza, were subtracted from this group since they were classified under the higher prevalence groups. After multiplying the estimate and CI by the group size, we predict that 109,088 (a range of 33,058 to 185,118) people are expected to develop PTSD.

### Group 6: Indirectly affected communities (living more than 80 km from the Gaza strip)

We identified two literature reviews relevant to this group. The first is the aforementioned review of Paz García-Vera, et al. (2016), which also examined PTSD prevalence in groups that were indirectly exposed to terror attacks (e.g., residents of cities that suffered terrorist attacks). We also relied on the review by Greene et al., (2018), specifically on studies that included comparison groups to the rocket exposure groups. The comparison groups included communities and cities within Israel that were exposed to low or no rocket attacks and, thus qualifying as indirectly affected.

#### Omission of studies

In the review by Paz García-Vera (2016), we omitted studies for the following reasons: six studies did not distinguish between people who were directly affected by the terror attack and those who were not; one study included help-seeking population; one study included a community not affected by the terror attack; and one study was conducted within a period of marked threat of terrorism. This resulted in only one study from this review being included in the meta-analysis. In the review by Green et al., (2018), 3 studies included a comparison group (i.e., low to no exposure to rocket attacks). All other studies did not include data of comparison groups and therefore were not considered for the current meta-analysis.

#### Meta-analysis of included studies

Four studies (with 5 samples) were included in the meta-analysis. The mean prevalence estimate was μ^ = 0.02 (95%CI 0.01-0.03). The Q-test and I2 statistic indicated that outcomes are heterogeneous and that a meaningful portion of the variance could be explained by true heterogeneity rather than chance (Q(5) = 143.52, p < 0.001, I2 = 97.34%, τ = .09).

#### Group size and numerical estimate and range

As the current population of the State of Israel is 9,767,718 (See Appendix 1), this group was calculated to be 3,433,286 (after subtraction of all other groups). After multiplying the estimate and CI by the group size, we predict that 70,714 (a range of 21,410 to 120,019) people are expected to develop PTSD. *Prevalence estimates which were not based on reviews and meta-analyses*

### Group 2: Close proximity to the October 7^th^ terror attacks

As mentioned, for two of the exposure groups we were not able to identify a relevant review. One of these groups was group 2 (close proximity to the October 7^th^ terror attacks), which was under severe threat of invasion by Hamas perpetrators. We searched for reviews specifically addressing a population that was not directly subject to a terror attack but was under perceived life threat due to close proximity to a terror event. We did not find any review that reflected such exposure. One relevant study was identified from the review by Paz García-Vera, et al., (2016) that was utilized for other groups. This study, conducted by North, et al., (2011), examined PTSD prevalence in direct survivors of terror attacks, as well as in employees of New York City companies affected by the events of September 11. The latter group was in close proximity to the terror attacks and experienced perceived life threats but did not personally experience the acts of terror. The prevalence estimate of this group was 0.21 (0.15 after adjustment with the correction factor). However, the size of this specific exposure group within the study was very small (N=19). Including only one study with such a small sample size can result in a biased estimate. Therefore, we chose to not rely on the estimate derived from this study. In the absence of other relevant studies, we used the estimates of group 4 (intensive rocket exposure), as civilians who were under threat of invasion by Hamas perpetrators were also under intense rocket attacks. We therefore relied on the estimate and confidence interval of group 4 (μ^= 0.10 (95% CI 0.03-0.17).

#### Group size and numerical estimate and range

This group amounted to a total of 121,061 people (After subtraction of group 1, see Appendix 1). After multiplying the aforementioned estimate and CI by the group size, we predict that 12,354 people (with a range of 3,744 to 20,964) are expected to develop PTSD.

### Group 5: Civilians under moderate exposure to rocket attacks (living 40-80 km from the Gaza Strip)

A second exposure group for which we could not identify a review was group 5 (moderate exposure to rocket attacks). The majority of studies regarding the exposure of a civilian population to rocket attacks was conducted in Israel, and included in the review by Greene et al., (2018) presented for group 4 (under intensive rocket attacks). However, this group shares similarities with both the intensive rocket exposure group, and the indirectly affected communities, which are characterized by low to no rocket exposure. Therefore, we addressed this group as representing an intermediate level of exposure between groups 4 and 6. Accordingly, to create the estimate for this group, we averaged the prevalence rates of groups 4 and 6. To create the range, we averaged the lower and upper bounds of the confidence intervals of groups 4 and 6. The computed estimate for this group was 0.06, and the range was 0.02 to 0.10.

#### Group size and numerical estimate and range

This group amounted to a total of 4,960,469 people (See Appendix 1). After multiplying the estimate and the range by the group size, we predict that 304,182 (a range of 92,166 to 516,198) people are expected to develop PTSD.

## Discussion

We developed a prediction model of the prevalence of PTSD following the October 7^th^ mass terrorist attack and the Israel-Hamas war for the varying levels of exposure across the Israeli population. Relying on meta-analyses and review papers, prevalence estimates ranged between 0.02 for the general population, defined as indirectly exposed to the events of the war, to 0.32 for those directly exposed to the October 7^th^ terror atrocities and the ensuant war. Considering the size of each exposure group, the model predicts that approximately 519,923 people (a range of 160,346 to 879,502) which constitute 5.3% of the Israeli population (a range of 1.6% to 9%), are expected to develop PTSD in the aftermath of the traumatic events.

A recent meta-analysis of PTSD prevalence in populations exposed to war and violent conflict, which included both clinical assessments and questionnaire-based assessments, indicated a 23.5% prevalence estimate in civilian populations (Lim, et al., 2022). Another meta-analysis of adult civilian war survivors who live in war-afflicted regions indicated a prevalence estimate of approximately 24% (Hoppen, et al., 2019). Our results present considerably smaller prevalence estimates compared to these studies. Importantly, studies regarding the mental health of the Israeli population following the current events have already indicated a high prevalence rate of PTSD and other post-traumatic sequelae of traumatic exposure. For example, a longitudinal survey in a representative sample (N= 710) of the adult population in Israel (aged 18-85), of whom 30 participants (4.3%) were directly exposed to the terror attacks, indicated an increase in the probable rate of PTSD, from 16% in August 2023 to 30% in November 2023, one month after the terror attacks (Levi-Belz et al., 2024).

This discrepancy might be accounted for the fact that the latter study included measurement of PTSD within the timeframe of the war, while our model reflects a prediction of PTSD in the war’s aftermath. Another broader methodological explanation concerns one of the inclusion criteria in the current model, that required the studies to assess PTSD specifically with regards to the exposure to war or terror. On the other hand, in the study by Levi-Belz et al. (2004), (similarly to several questionnaire-based assessments of PTSD prevalence), participants were instructed to consider any traumatic event they can recall (and not a specific traumatic exposure related to war or terror). Such measurements may inflate PTSD prevalence estimates compared to studies included in our model. Finally, we intentionally implemented a correction factor for questionnaire-based measurements in order to avoid inflation of estimates. This might also account for the lower expected PTSD rates presented in our model.

While this review focused on the Israeli population, it is imperative to note that the implications for Palestinians living in Gaza have been widespread and severe, affecting both those directly involved and those uninvolved, with mass casualties and displacement of the civilian population. It is important for future research to also estimate the prevalence of PTSD among Palestinian civilians in Gaza, which requires access to relevant sources of information.

### Limitations

While this model focused on predicting PTSD, other forms of psychopathology are also highly prevalent after exposure to war and terror, particularly depression and anxiety disorders (e.g., Hoppen, et al., 2021). Moreover, war exposure is associated with additional stressors that do not include direct exposure to violence, such as financial, occupational, and interpersonal challenges. These are also expected to be associated with the extent of distress and mental health outcomes (even if not specifically to PTSD).

In addition, given the ecological, population-wide approach, we could not address specific personal and sub-population risk and protective factors that were found to be associated with the development of PTSD following exposure to terrorist attacks and war atrocities, among others: prior psychopathology alcohol/substance abuse disorders, gender, socio-economic and marital status (Shalev, et al., 2019; Sayed, et al., 2015). Nonetheless, the vast majority of research presented in the reviews included population-based samples, which are considered to be representative of the general population and do not exclude individuals who have prior risk factors. Therefore, we assume that this issue does not substantially compromise the validity of our findings.

Some specific subgroups were not included in this model (e.g., aid teams and individuals who have a relative who was killed, injured, or abducted during the war). Studies have shown that these types of exposure are associated with an increased risk for PTSD (Chiu et al, 2011; Cukor, et al., 2011, for aid teams; North, et al., 2011; Miguel-Tobal, et al., 2005, for relatives of victims). However, it was unfeasible to assess group size and estimate the unique prevalence rates of such forms of exposure; therefore, they were not included in the final model. Similarly, approximately 200,000 Israelis were evacuated from their homes (the majority of them from the southern area, and others from the communities close to the Israel-Lebanon border; Peleg & Gendelman, 2023), and they have an increased risk of detrimental mental health outcomes (Husain, et al., 2011). However, we did not address displacement as exposure to terror since some of the displacements occurred for preventive purposes. Moreover, many of those displaced belong to exposure groups 1, 2, and 4, which were all subject to direct terror or rocket attacks.

This study was conducted while the Israel-Hamas war was still going on. Although we reviewed research that examined PTSD prevalence in the aftermath of terror attacks and war exposures, large groups of the Israeli population are still experiencing traumatic events, and the number of casualties and the death toll are rising and undetermined. We relied on the most validated information within our reach, with a preference for government-originated information, yet true group sizes might change to some degree.

Although we attempted to identify relevant reviews for each exposure group in the most nuanced manner possible, the singular characteristics of the October 7^th^ terror attack and the Israel-Hamas war might compromise our ability to rely on and extrapolate from previous research for the purpose of predicting PTSD in the current situation. Hoffman et al., (2011) study regarding the unique elements of ongoing trauma is particularly relevant to war and terror exposure in the Israeli population. Israeli citizens have been exposed to various forms of terror and war ever since the establishment of the state. Many of the studies presented in this review did not address such unique long-term exposure and have referred to traumatic events that include a specific, often single-time terror exposure (e.g., Paz Garcia-Vera, et al., 2016).

Along with this notion, we have several reasons to assume that this model can be considered conservative with regards to its prediction of PTSD prevalence. Firstly, we did not account for some risk factors that do not qualify as direct exposure to terror. One such risk factor is the intense exposure to viral media coverage of the atrocities, which included videos and testimonies of extreme violence, torture, and sexual assault. Indirect exposure to trauma through the media has been shown to relate to post-traumatic symptoms (e.g., Neria & Sullivan, 2011). In the absence of specific data regarding the extent of media exposure across the population, and due to the challenge of assessing the effects of such exposure on PTSD prevalence, this factor was not accounted for in the literature review process. Nonetheless, media exposure is considered to be widespread across all exposure groups, and there is reason to expect that prevalence rates might be higher due to this form of exposure.

Another limitation of our model is that we did not account for a possible additive risk for PTSD due to more than one form of exposure. As mentioned, in cases of multifaceted exposure, we chose the highest prevalence rate. This decision does not account for a possible additive component of several types of terror exposure. Additionally, for group 2 (close proximity to the October 7^th^ attack), we relied on less severe exposure (rocket attacks) compared to the actual exposure.

We presented several reasons to assume our model might underestimate the prevalence of PTSD, yet some of the methodological decisions that we made might result in an overestimation of prevalence rates. Specifically, in longitudinal studies that included more than one prevalence estimate we chose the highest prevalence estimate. This is because we wanted to capture the extent to which individuals within each group are expected to develop PTSD, without addressing a specific time frame. As most of the studies presented in the review showed a general trend of decline in PTSD prevalence in longitudinal examinations, this might result in an overestimation of the PTSD prevalence, and not account for some of the natural processes of decline in the magnitude of post-traumatic symptoms. It is also important to note that the high heterogeneity in the timing of PTSD measurement within the reviewed studies compromises the precision of the model in terms of time-specific prediction and does not allow for predicting PTSD prevalence at a specific time period after the war exposure.

There are two additional limitations that might not over or underestimate the prevalence estimates but are important to mention. The first is the considerable heterogeneity within exposure groups, particularly in the exposure to rockets groups. Within the same exposure groups, some civilians were much more exposed to rocket attacks than others, even when distances between their areas of residency were small. The second is the fact that we did not differentiate between children and adults, due to the ambiguity of findings regarding differences between children and adults in PTSD prevalence after exposure to trauma.

Another limitation that is not expected to significantly change prevalence estimates but has important clinical implications is the fact that subgroups of exposure groups 1 (civilian direct victims) and 3 (soldiers) were subject to extremely severe traumas, including abduction, torture, or sexual assault. These subgroups are small in magnitude compared to the groups in which they are included. However, these subgroups are at much higher risk for PTSD than the estimates of their respective groups. For example, the review by Johnson and Thompson (2008) regarding torture survivors presented several studies reporting more than 50% PTSD prevalence.

One aspect that we did not cover in this study is the severity of PTSD symptoms. While using a dichotomous cutoff is common in clinical and epidemiological research, it is important to acknowledge that PTSD, like other mental health disorders, exhibits significant variability in terms of the severity and intensity of its clinical presentation. This variability holds important implications, particularly in the context of implementing research findings for intervention planning, and future research would benefit from further exploration of this topic.

### Policy and Clinical Implications

Given the prediction of hundreds of thousands of people suffering from PTSD in Israel, stepped, multi-layered, monitored, and cost-effective mental health services should be planned to reach the maximum number of individuals suffering from PTSD or other trauma-related mental disorders. Although most of the exposed population will recover from PTSD without the need for psychological or psychiatric intervention (Rosellini, et al., 2018), immediate, timely, and comprehensive interventions are required to prevent chronic and severe morbidity for the population at risk. This model can help policymakers, philanthropists, researchers, and clinicians in planning future steps and preparing adequate short and long-term responses for the mental health needs of the population exposed to mass trauma. Additionally, the underpinning of the model can be utilized, with relevant, necessary modifications, in other occasions of mass-trauma exposure.

When applying this model’s results to the public Israeli mental health services, it is crucial to acknowledge the pre-existing significant deficits in these services prior to the recent conflict. The wait times for services extended egregiously, often surpassing five months following an initial referral, coupled with a stark insufficiency of professionals, including psychiatrists and psychotherapists, within the public sector. This shortfall was unable to meet the population’s demands (Israel State Comptroller Annual Report, 2020). Given these pronounced deficiencies, the public mental health system is anticipated to encounter considerable obstacles in delivering interventions commensurate with the post-conflict expected needs of the population. Furthermore, the prevalent reliance on individual psychotherapy across various mental health services in Israel appears ill-suited to manage the anticipated breadth of the mental health crisis effectively.

Consequently, there is a pressing need for the adoption of comprehensive, system-wide models facilitating large-scale interventions. Such models should incorporate evidence-based group therapies (Schwartze et al., 2017) and short-term individual protocols (Jensen et al., 2014), initiatives for prevention and early intervention (Birur et al., 2017), the utilization of digital technologies for monitoring and management of mental health symptoms (Wang et al., 2018), and the training of lay psychological counselors among community workers, as addressed by the World Health Organization (2022). There is also critical importance in monitoring of services and outcomes (Clark et al, 2018). Moreover, policymakers might also consider the allocation of differential funding contingent upon the services’ capacity to enact such expansive and large-scale interventions.

## CONCLUSIONS

The expected number of PTSD cases following the October 7^th^ events and the Israel-Hamas war is expected to be substantial. Strategic planning and implementation of large-scale interventions represent a potential avenue to bridge the disparity between the extensive needs of the population and the mental health system’s limited resources.

## Declarations

### Authors’ contributions

DK has designed the research, collected and reviewed data, conducted the inclusion and omission of studies, conducted meta-analyses and written the article.

MG has initiated the project, designed the research, collected and reviewed data, and conducted the inclusion and omission of studies.

YN has provided critical revisions of the article.

IL and AS have contributed substantially to the design of the research, participated in generating inclusion and exclusion criteria for meta-analyses, and provided critical revisions of the article.

### Availability of data and materials

No original data was included in this study. All articles included in meta-analyses are addressed in the manuscript and supplementary material.

## Acknowledgments

The authors wish to thank Ariel Knafo-Noam and Yaakov Greenwald for their helpful comments regarding the manuscript, Gidi Kadosh for raising the need and his help in the conceptualization of the project, Chen Pizanti for her assistance in the collection of data, and Effective Altruism Israel for the support of the project.

## Ethics approval and consent to participate

Not applicable. The present study relied on meta-analyses of prevalence rates presented in previously published articles and does not include any additional or original data. The data extracted from each article included prevalence mean and group size, information that was openly available in all articles.

## Consent for publication

Not applicable.

---

**Appendix 1.**
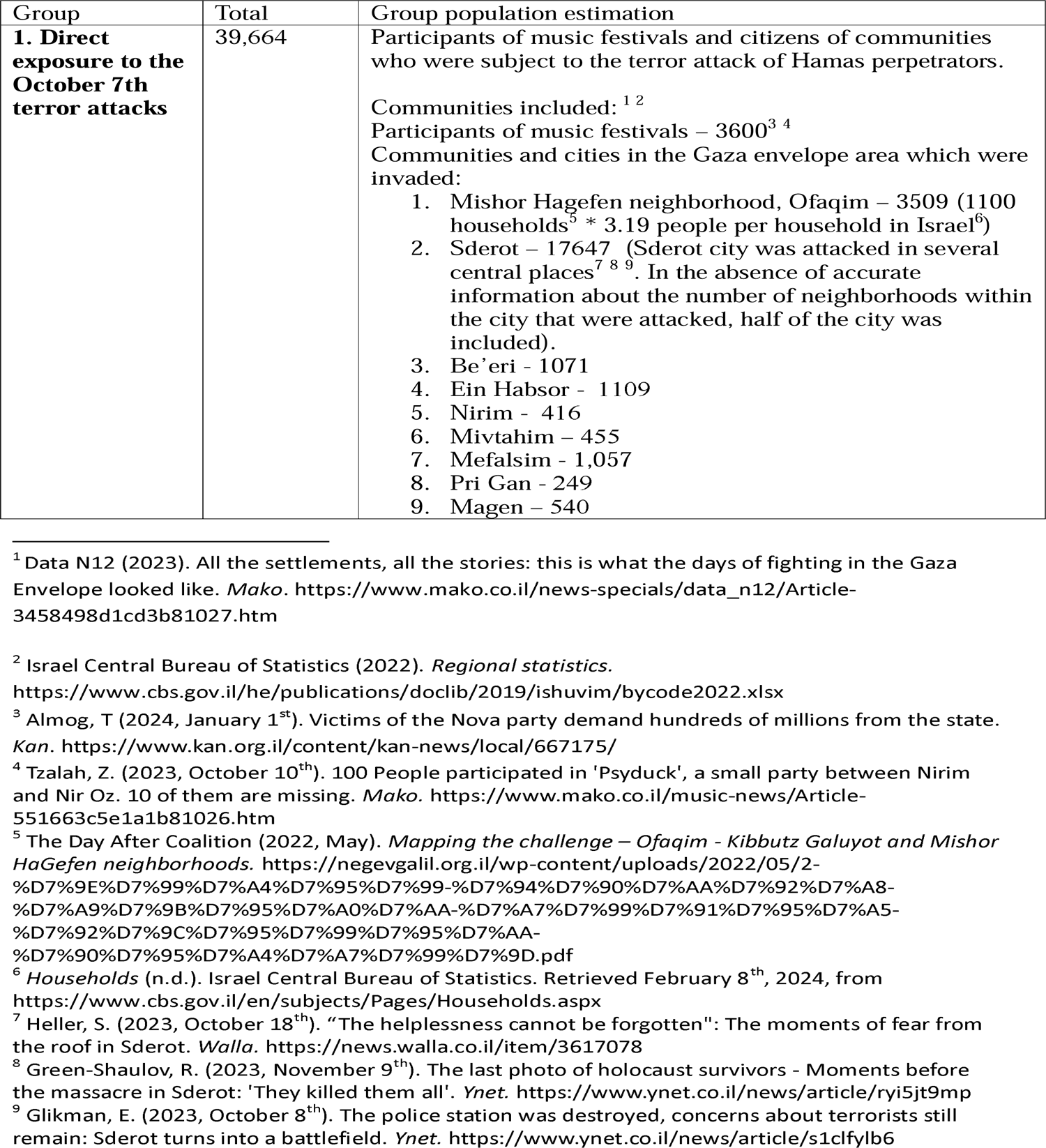
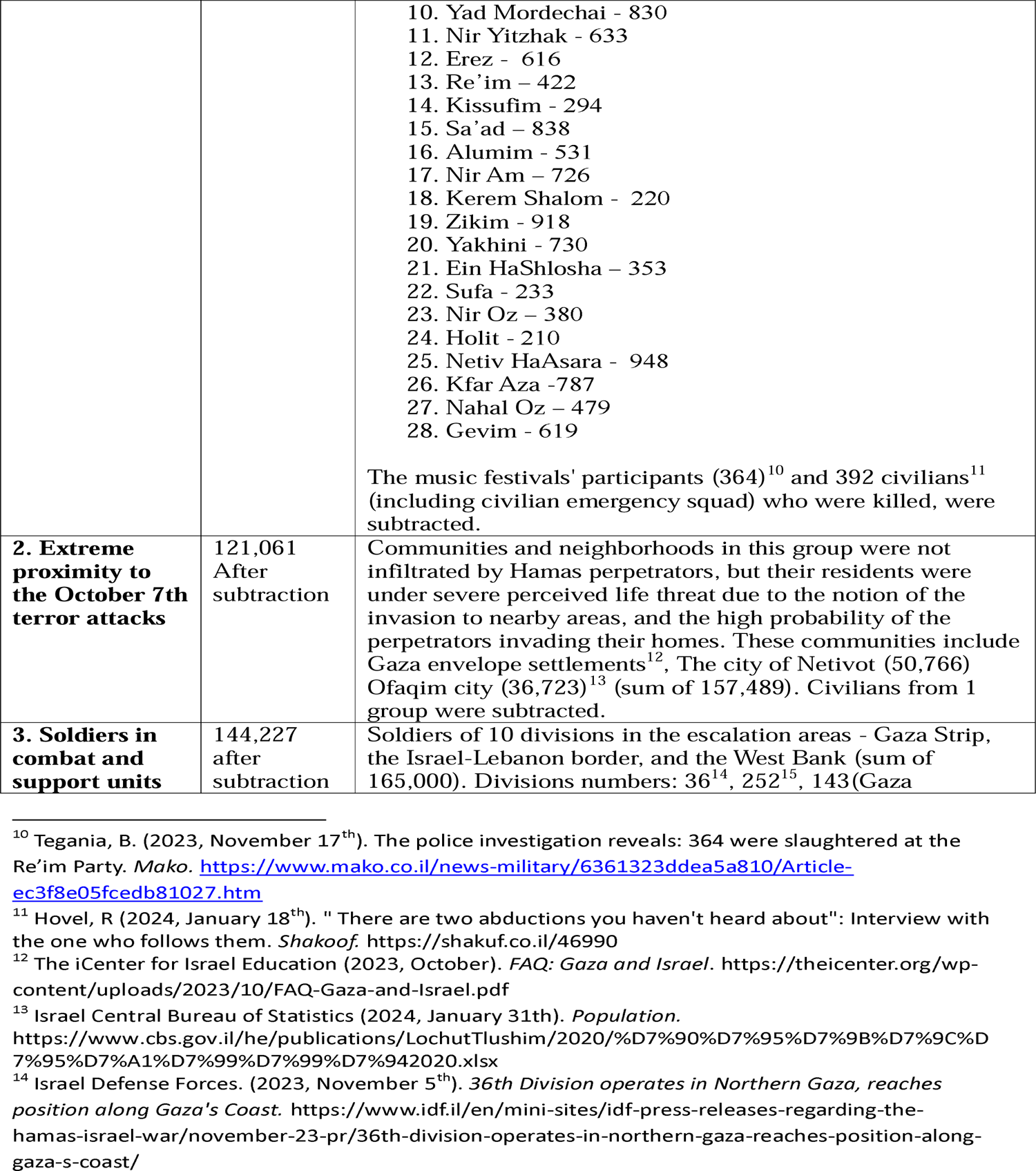
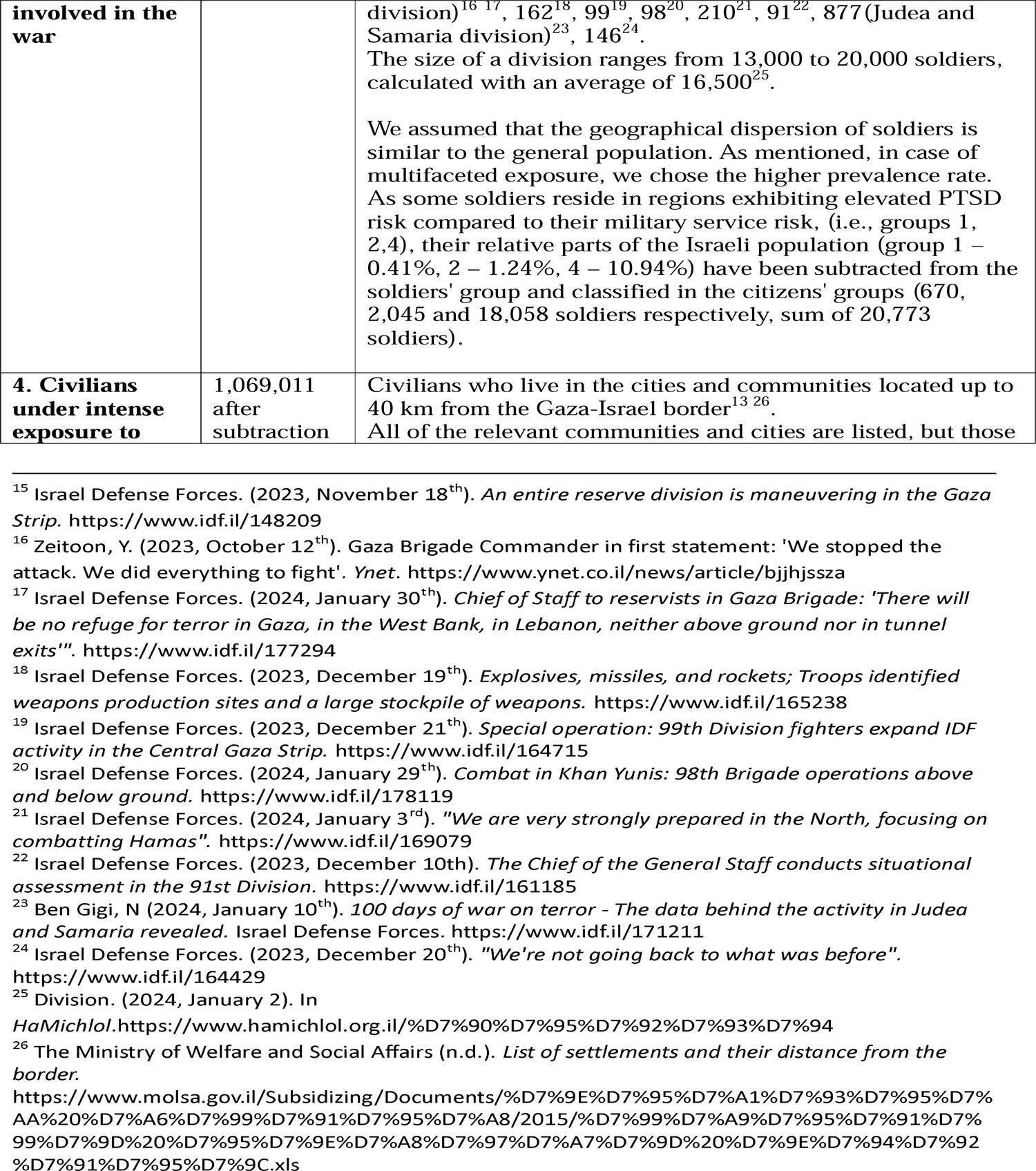
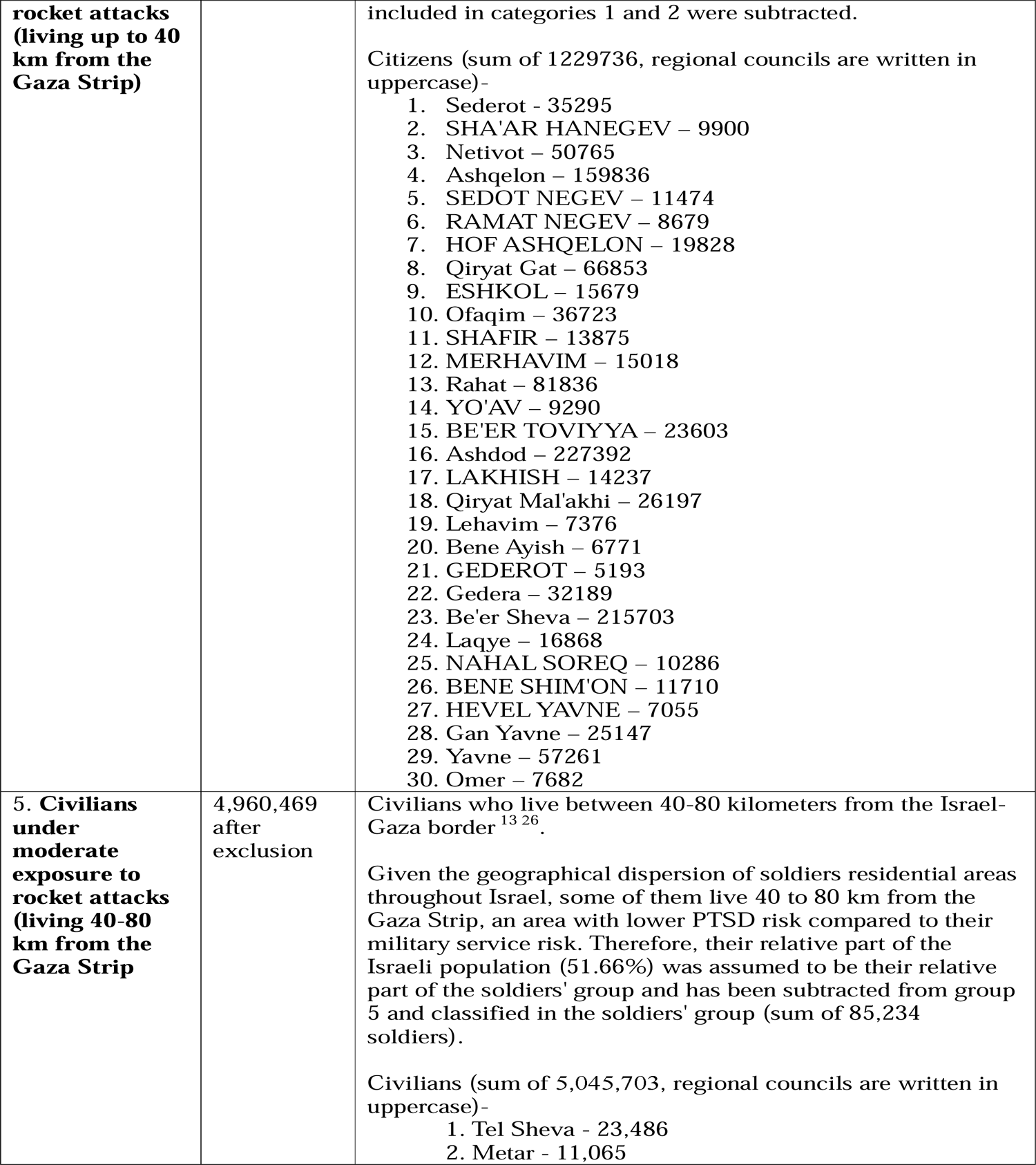
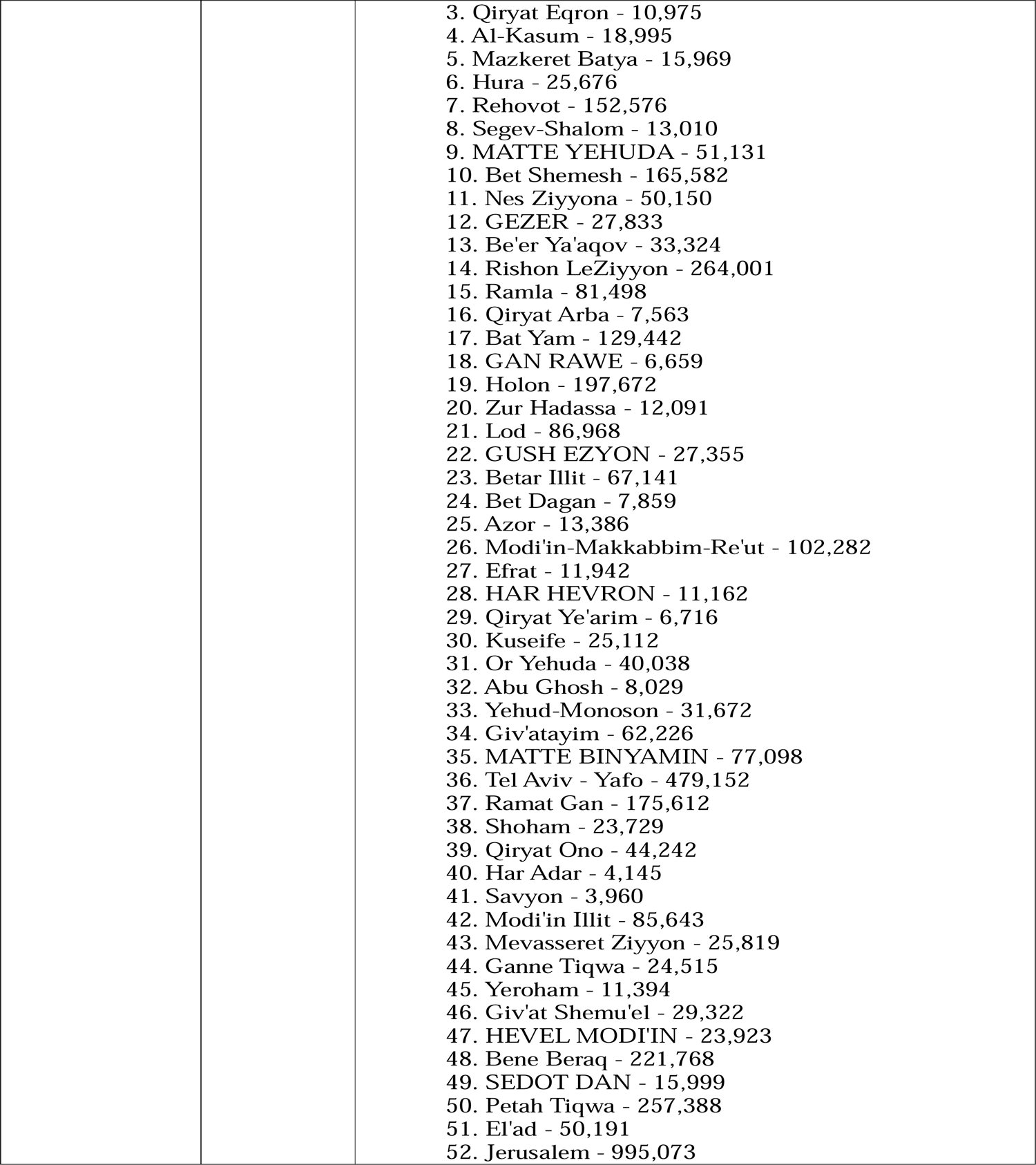
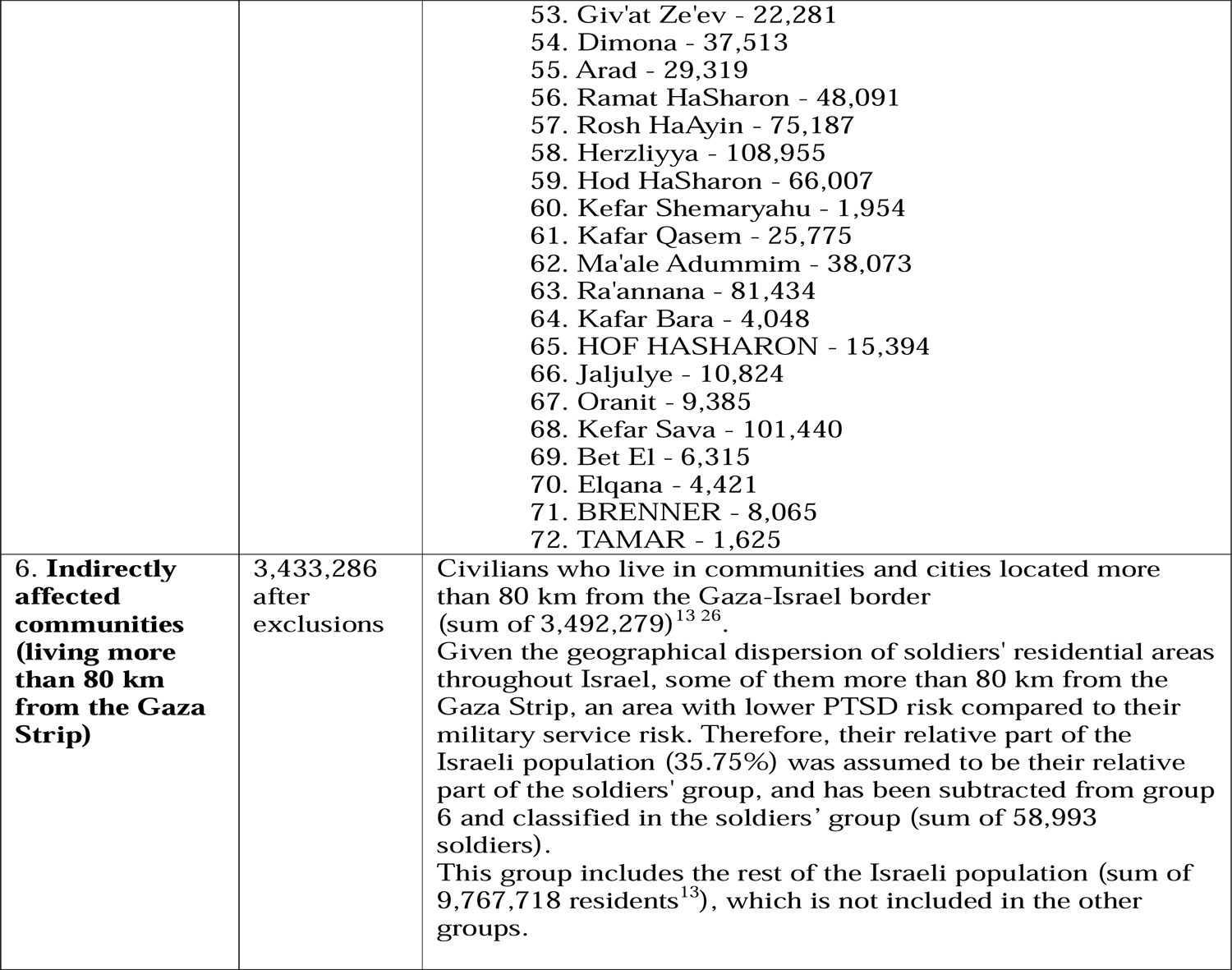
Numerical estimations for exposure groups.

