## Supplementary material for "A Prediction Model of PTSD in the Israeli Population in the Aftermath of October 7^th^, 2023, Terrorist Attack and the Israel-Hamas War"

**Omission of studies in meta-analyses**

**Group 1: Direct exposure to the October 7th terror attacks**.

In the systematic review by Paz García-Vera, et al. (2016), one study (Ankri, et al., 2010) was omitted as it included help-seeking population.

**Group 3: Soldiers in combat and support units involved in the war.**

In the review by Richardson, et al. (2010) studies were omitted for the following reasons: 18 studies included questionnaire assessments or non-clinical assessments (Barrett et al., 2002; Browne et al., 2007; Dohrenwend et al., 2006; Eisen et al., 2004; Goldberg et al., 1990; Goss-Gilroy 1998; Gray et al., 2002; Hoge et al., 2004; Hoge et al., 2006; Hotopf et al., 2006; Iowa Persian Gulf Study Group, 1997; Iversen et al., 2008; Jones et al., 2006; Kang et al., 2003; Smith et al., 2008; Stretch et al., 1993; Thompson et al., 2006; Unwin et al., 1999; Wolfe et al., 1999); two studies re-analyzed existing data from another sample (Dohrenwend et al., 2006; Thompson et al., 2006); one study included help-seeking population (Lee et al., 2002); and one study that did not examine current PTSD prevalence (Ikin, et al., 2004).

In the review by Xue, et al., (2015), studies were omitted for the following reasons: 23 studies included questionnaire assessments or non-clinical assessments (Barrett et al., 2002; Booth-Kewleyet al., 2010; Du Preez et al., 2011; Goldmann et al., 2012; Harbertson et al., 2013; Iversen et al., 2008; Iversen et al., 2009; Jones et al., 2006; Jones et al., 2012; Kline et al., 2013; Koenen et al., 2003; LeardMann et al., 2009; LeardMann et al., 2010; Macera et al., 2014; MacGregor et al., 2013; McCarren et al., 1995; Phillips et al., 2010; Riviere et al., 2011; Rona et al., 2007; Rona et al., 2012; Sandweiss et al., 2011; Van Liempt et al., 2013; Wells et al., 2012); three studies examined PTSD diagnosis in medical files without specific reference to the war exposure (Mayo, et al., 2013; Maguen, et al., 2012; MacGregor, et al., 2012); and three studies did not examine current prevalence (Tracie, et al., 2013; Dohrenwend, et al., 2008; Koenen, et al., 2002).

**Group 4: Civilians under intense exposure to rocket attacks (living up to 40 km from the Gaza Strip)**.

In the review by Greene, et al., 2018, out of the twelve studies that presented included PTSD prevalence estimates, studies were omitted for the following reasons: two studies examined PTSD within the timeframe of war exposure (Besser and Neria, 2010; Nuttman-Shwartz, et al., 2015); one study did not specify whether measurement was conducted during or after war exposure (Nuttman-Shwart, et al., 2014); and two studies did not clarify whether they assessed PTSD symptoms that related specifically to rocket attacks (Ben-Ezra et al., 2015; Stein, et al., 2013).

**Group 6: Indirectly affected communities** (**living more than 80 km from the Gaza strip)**.

In the review by Paz García-Vera (2016), studies were omitted for the following reasons: six studies did not distinguish between people who were directly affected by the terror attack and those who were not (Miguel-Tobal, et al., 2005; Nandi, et al., 2005; Lawyer, et al., 2006; Galea, et al., 2004; Boscarino, et al., 2004; Stuber, et al., 2006); one study included help-seeking population (Shear, et al., 2006); one study included a community not affected by the terror attack (Henrikson, et al., 2010); and one study was conducted within a period of marked threat of terrorism (Hobfall, et al., 2011).

**Forest plots of meta-analyses**

Direct exposure to terror attacks meta-analysis

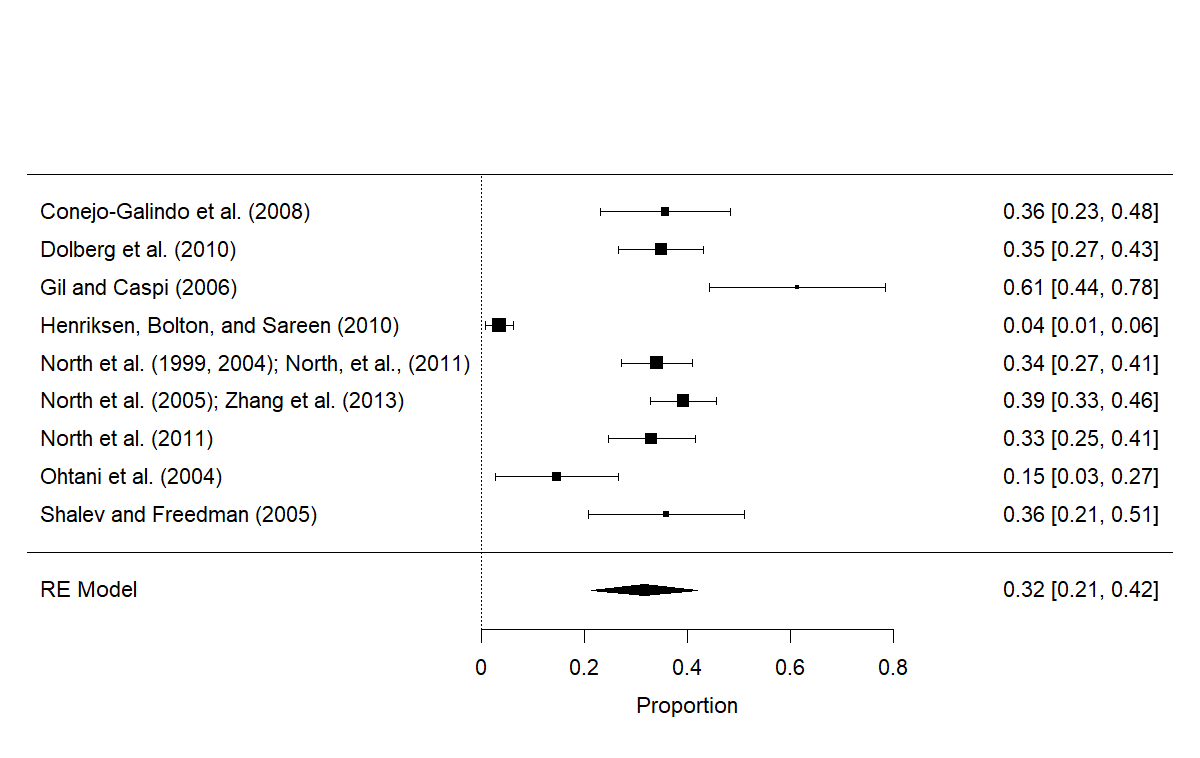

Soldiers meta-analysis

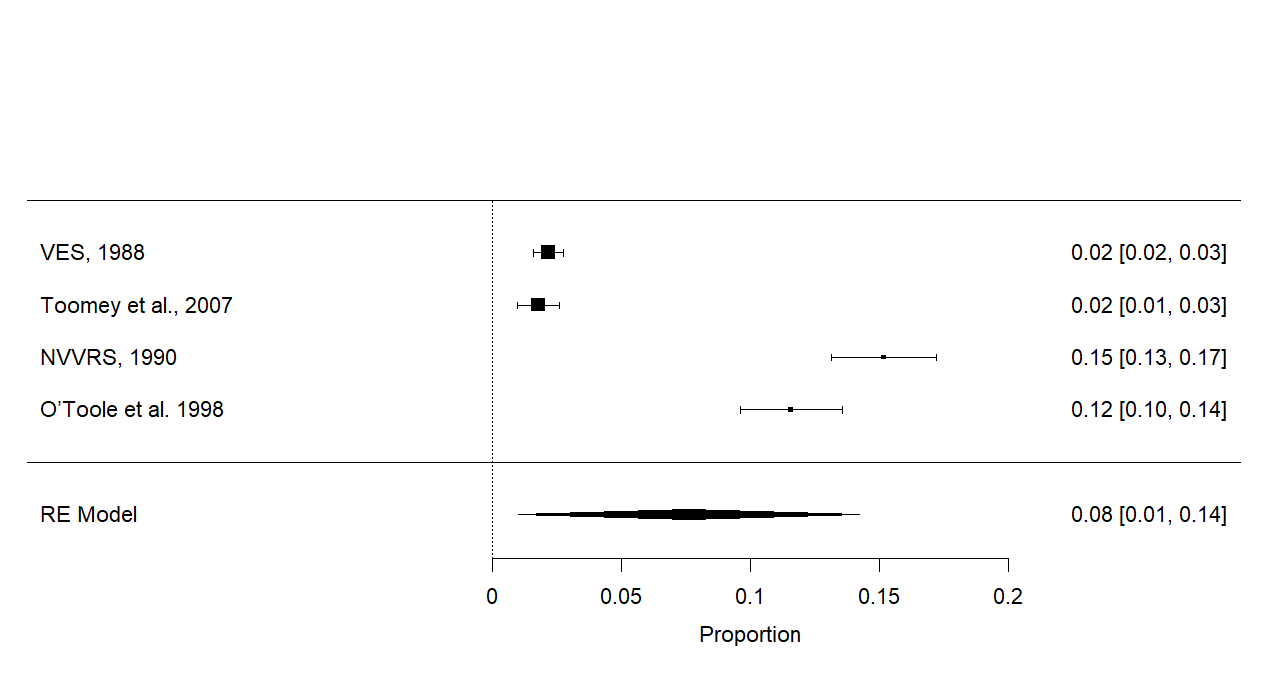

Intense rocket exposure meta-analysis

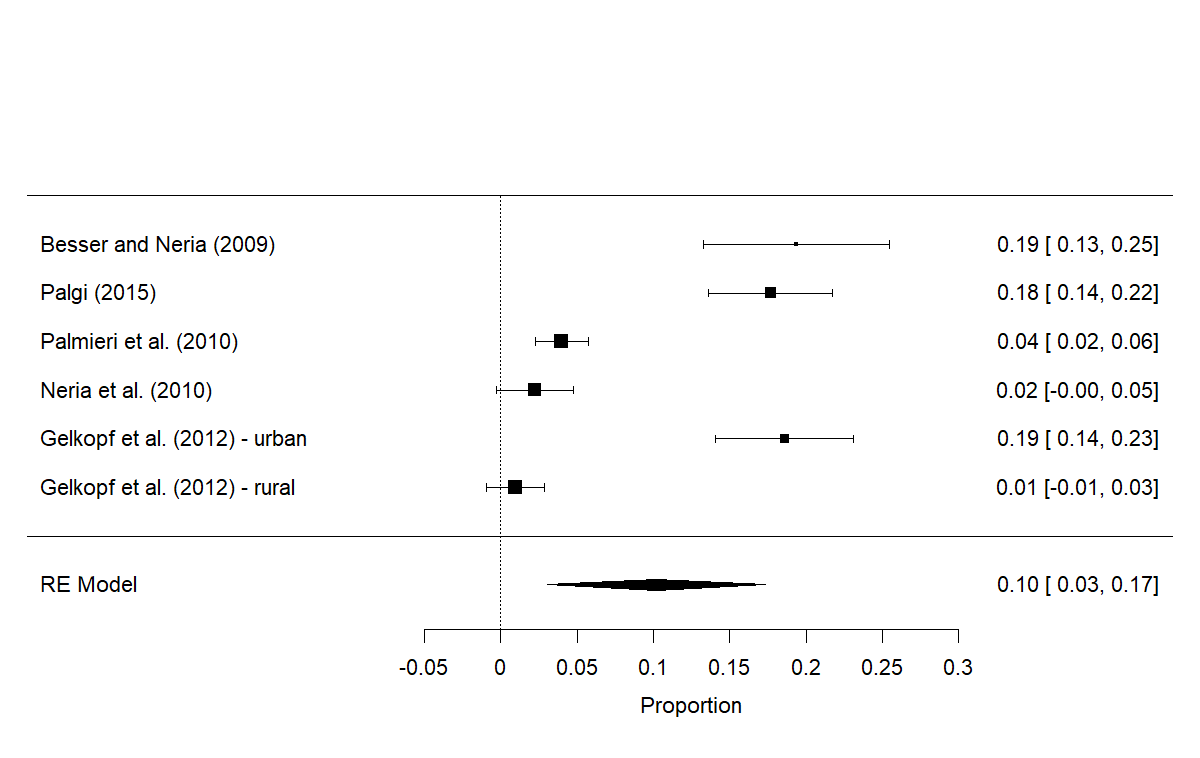

Indirectly affected community meta-analysis

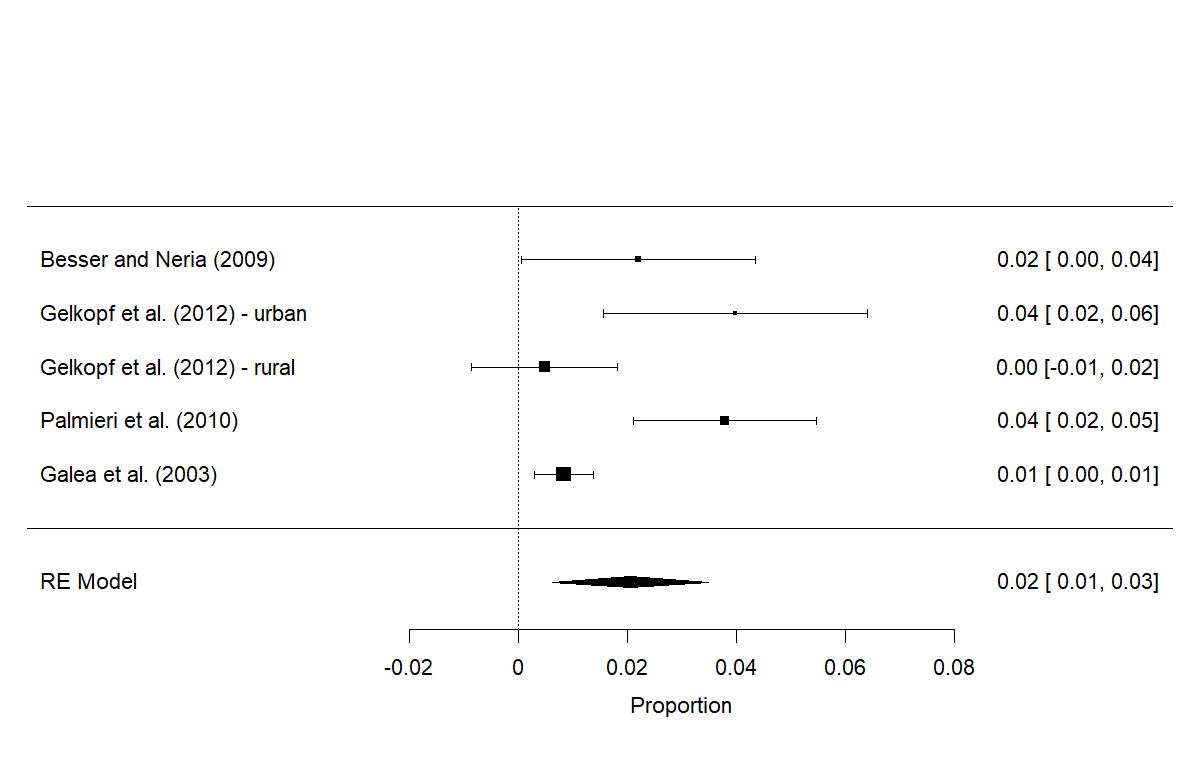

**REFRENCES**

Ankri, Y. L., Bachar, E., & Shalev, A. Y. (2010). Reactions to terror attacks in ultra-Orthodox Jews: The cost of maintaining strict identity. *Psychiatry: Interpersonal and Biological Processes*, 73(2), 190-197.

Barrett, D. H., Doebbeling, C. C., Schwartz, D. A., Voelker, M. D., Falter, K. H., Woolson, R. F., & Doebbeling, B. N. (2002). Posttraumatic stress disorder and self-reported physical health status among US Military personnel serving during the Gulf War period: a population-based study. *Psychosomatics*, 43(3), 195-205.

Barrett, D. H., Doebbeling, C. C., Schwartz, D. A., Voelker, M. D., Falter, K. H., Woolson, R. F., & Doebbeling, B. N. (2002). Posttraumatic stress disorder and self-reported physical health status among US Military personnel serving during the Gulf War period: a population-based study. *Psychosomatics*, 43(3), 195-205.‏

Besser, A., & Neria, Y. (2009). PTSD symptoms, satisfaction with life, and prejudicial attitudes toward the adversary among Israeli civilians exposed to ongoing missile attacks. *Journal of Traumatic Stress, 22*, 268 –275.

Besser, A., & Neria, Y. (2010). The effects of insecure attachment orientations and perceived social support on posttraumatic stress and depressive symptoms among civilians exposed to the 2009 Israel–Gaza war: A follow-up Cross-Lagged panel design study. *Journal of Research in Personality*, 44(3), 335-341.

Booth‐Kewley, S., Larson, G. E., Highfill‐McRoy, R. M., Garland, C. F., & Gaskin, T. A. (2010). Correlates of posttraumatic stress disorder symptoms in Marines back from war. Journal of Traumatic Stress: Official Publication of The International Society for Traumatic Stress Studies, 23(1), 69-77.‏

Boscarino, J. A., Adams, R. E., & Figley, C. R. (2004). Mental health service use 1-year after the world trade center disaster: Implications for mental health care*. General Hospital Psychiatry*, 26, 346–358

Browne, T., Hull, L., Horn, O., Jones, M., Murphy, D., Fear, N. T., ... & Hotopf, M. (2007). Explanations for the increase in mental health problems in UK reserve forces who have served in Iraq. *The British journal of psychiatry*, 190(6), 484-489.

Centers for Disease Control (1988). Vietnam Experience Study: health status of Vietnam veterans: psychosocial characteristics. *JAMA*, *259,* 2701–2707. [VES]Conejo-Galindo, J., Medina, O., Fraguas, D., Tera´n, S., Sainz-Corto´n, E., & Arango, C. (2008). Psychopathological sequelae of the 11 March terrorist attacks in Madrid: An epidemiological study of victims treated in a hospital. *European Archives of Psychiatry and Clinical Neuroscience, 258*, 28–34.

Dohrenwend, B. P., Turner, J. B., Turse, N. A., Adams, B. G., Koenen, K. C., & Marshall, R. (2006). The psychological risks of Vietnam for US veterans: a revisit with new data and methods. *Science*, 313(5789), 979-982.

Dohrenwend, B. P., Turner, J. B., Turse, N. A., Lewis‐Fernandez, R., & Yager, T. J. (2008). War‐related posttraumatic stress disorder in Black, Hispanic, and majority White Vietnam veterans: The roles of exposure and vulnerability. *Journal of Traumatic Stress*: Official Publication of The International Society for Traumatic Stress Studies, 21(2), 133-141.

Dolberg, O. T., Barkai, G., Leor, A., Rapoport, H., Bloch, M., & Schreiber, S. (2010). Injured civilian survivors of suicide bomb attacks: From partial PTSD to recovery or to traumatisation. Where is the turning point*? The World Journal of Biological Psychiatry, 11*, 344–351.

Du Preez, J., Sundin, J., Wessely, S., & Fear, N. T. (2012). Unit cohesion and mental health in the UK armed forces. *Occupational medicine*, 62(1), 47-53.

Du Preez, J., Sundin, J., Wessely, S., & Fear, N. T. (2012). Unit cohesion and mental health in the UK armed forces. *Occupational medicine*, 62(1), 47-53.‏

Eisen, S. A., Griffith, K. H., Xian, H., Scherrer, J. F., Fischer, I. D., Chantarujikapong, S., Hunter, J., True, W. R., Lyons, M. J. & Tsuang, M. T. (2004). Lifetime and 12-month prevalence of psychiatric disorders in 8,169 male Vietnam War era veterans. *Military medicine*, 169(11), 896-902.

Galea, S., Vlahov, D., Resnick, H., Ahern, J., Susser, E., Gold, J., ... & Kilpatrick, D. (2003). Trends of probable post-traumatic stress disorder in New York City after the September 11 terrorist attacks. *American journal of epidemiology*, *158*(6), 514-524.

Galea, S., Vlahov, D., Tracy, M., Hoover, D. R., Resnick, H., & Kilpatrick, D. (2004). Hispanic ethnicity and post-traumatic stress disorder after a disaster: Evidence from a general population survey after September 11, 2001. *Annals of Epidemiology*, 14, 520–531.

Gelkopf, M., Berger, R., Bleich, A., & Silver, R. C. (2012). Protective factors and predictors of vulnerability to chronic stress: A comparative study of 4 communities after 7 years of continuous rocket fire. *Social Science & Medicine, 74*, 757–766.

Gil, S., & Caspi, Y. (2006). Personality traits, coping style, and perceived threat as predictors of posttraumatic stress disorder after exposure to a terrorist attack: A prospective study. *Psychosomatic Medicine, 68*, 904–909.

Greene, T., Itzhaky, L., Bronstein, I., & Solomon, Z. (2018). Psychopathology, risk, and resilience under exposure to continuous traumatic stress: A systematic review of studies among adults living in southern Israel. *Traumatology*, *24*(2), 83.‏

Goldberg, J., Eisen, S. A., True, W. R., & Henderson, W. G. (1990). A twin study of the effects of the Vietnam conflict on alcohol drinking patterns. *American Journal of Public Health*, 80(5), 570-574.

Goldmann, E., Calabrese, J. R., Prescott, M. R., Tamburrino, M., Liberzon, I., Slembarski, R., ... & Galea, S. (2012). Potentially modifiable pre-, peri-, and postdeployment characteristics associated with deployment-related posttraumatic stress disorder among ohio army national guard soldiers. *Annals of epidemiology*, 22(2), 71-78.‏

Harbertson, J., Grillo, M., Zimulinda, E., Murego, C., Cronan, T., May, S., ... & Shaffer, R. (2013). Prevalence of PTSD and depression, and associated sexual risk factors, among male R wanda D efense F orces military personnel. *Tropical Medicine & International Health*, 18(8), 925-933.‏

Harbertson, J., Grillo, M., Zimulinda, E., Murego, C., Cronan, T., May, S., Brodine, S., Sebagabo, M., Araneta, M. R. G. & Shaffer, R. (2013). Prevalence of PTSD and depression, and associated sexual risk factors, among male Rwanda Defense Forces military personnel. *Tropical Medicine & International Health*, 18(8), 925-933.

Henriksen, C. A., Bolton, J. M., & Sareen, J. (2010). The psychological impact of terrorist attacks: Examining a dose-response relationship between exposure to 9/11 and axis I mental disorders. *Depression and Anxiety*, 27, 993–1000

Hobfoll, S. E., Canetti, D., Hall, B. J., Brom, D., Palmieri, P. A., Johnson, R. J., ... Galea, S. (2011). Are community studies of psychological trauma’s impact accurate? A study among Jews and Palestinians. *Psychological Assessment*, 23, 599–605.

Hoge, C. W., Auchterlonie, J. L., & Milliken, C. S. (2006). Mental health problems, use of mental health services, and attrition from military service after returning from deployment to Iraq or Afghanistan. *Jama*, 295(9), 1023-1032.

Hoge, C. W., Castro, C. A., Messer, S. C., McGurk, D., Cotting, D. I., & Koffman, R. L. (2004). Combat duty in Iraq and Afghanistan, mental health problems, and barriers to care. *New England journal of medicine*, 351(1), 13-22

Hotopf, M., Hull, L., Fear, N. T., Browne, T., Horn, O., Iversen, A., ... & Wessely, S. (2006). The health of UK military personnel who deployed to the 2003 Iraq war: a cohort study. *The lancet*, 367(9524), 1731-1741.

Ikin, J. F., Sim, M. R., Creamer, M. C., Forbes, A. B., McKenzie, D. P., Kelsall, H. L., Glass, D. C., McFarlane, A. C., Abramson, M. J., Ittak, P., Dwyer, T., Blizzard, L., Delaney, K. R., Horsley, K. W. A., Harrex, W. K.& Schwarz, H. (2004). War-related psychological stressors and risk of psychological disorders in Australian veterans of the 1991 Gulf War. *The British Journal of Psychiatry*, 185(2), 116-126.

Iversen, A. C., Fear, N. T., Ehlers, A., Hughes, J. H., Hull, L., Earnshaw, M., ... & Hotopf, M. (2008). Risk factors for post-traumatic stress disorder among UK Armed Forces personnel. *Psychological medicine*, 38(4), 511-522.‏

Iversen, A. C., Fear, N. T., Ehlers, A., Hughes, J. H., Hull, L., Earnshaw, M., Greenberg, N., Rona, R., Wessely, S. & Hotopf, M. (2008). Risk factors for post-traumatic stress disorder among UK Armed Forces personnel. *Psychological medicine*, 38(4), 511-522.

Iversen, A. C., van Staden, L., Hughes, J. H., Browne, T., Hull, L., Hall, J., ... & Fear, N. T. (2009). The prevalence of common mental disorders and PTSD in the UK military: using data from a clinical interview-based study. *BMC psychiatry*, 9(1), 1-12.‏

Jones, M., Rona, R. J., Hooper, R., & Wesseley, S. (2006). The burden of psychological symptoms in UK Armed Forces. *Occupational medicine*, 56(5), 322-328.

Jones, M., Rona, R. J., Hooper, R., & Wesseley, S. (2006). The burden of psychological symptoms in UK Armed Forces. *Occupational medicine*, 56(5), 322-328.‏

Jones, M., Sundin, J., Goodwin, L., Hull, L., Fear, N. T., Wessely, S., & Rona, R. J. (2013). What explains post-traumatic stress disorder (PTSD) in UK service personnel: deployment or something else?. *Psychological medicine*, 43(8), 1703-1712.‏

Kang, H. K., Natelson, B. H., Mahan, C. M., Lee, K. Y., & Murphy, F. M. (2003). Post-traumatic stress disorder and chronic fatigue syndrome-like illness among Gulf War veterans: a population-based survey of 30,000 veterans. *American journal of epidemiology*, 157(2), 141-148.

Kline, A., Ciccone, D. S., Weiner, M., Interian, A., St. Hill, L., Falca-Dodson, M., ... & Losonczy, M. (2013). Gender differences in the risk and protective factors associated with PTSD: a prospective study of National Guard troops deployed to Iraq. *Psychiatry: interpersonal & biological processes*, 76(3), 256-272.‏

Koenen, K. C., Harley, R., Lyons, M. J., Wolfe, J., Simpson, J. C., Goldberg, J., Eisen, S. A. & Tsuang, M. (2002). A twin registry study of familial and individual risk factors for trauma exposure and posttraumatic stress disorder. *The Journal of nervous and mental disease*, 190(4), 209-218.

Koenen, K. C., Stellman, J. M., Stellman, S. D., & Sommer Jr, J. F. (2003). Risk factors for course of posttraumatic stress disorder among Vietnam veterans: a 14-year follow-up of American Legionnaires*. Journal of consulting and clinical psychology*, 71(6), 980.‏

Kulka, R. A., Schlenger, W.E., Fairbank, J.A., et al (1990). Trauma and the Vietnam War generation: report of findings from the National Vietnam Veterans Readjustment Study. Brunner/ Mazel. [NVVRS]

Lawyer, S. R., Resnick, H. S., Galea, S., Ahern, J., Kilpatrick, D. G., & Vlahov, D. (2006). Predictors of peritraumatic reactions and PTSD following the September 11th terrorist attacks. *Psychiatry*, 69, 130–141

LeardMann, C. A., Smith, B., & Ryan, M. A. (2010). Do adverse childhood experiences increase the risk of postdeployment posttraumatic stress disorder in US Marines?. *BMC Public Health*, 10(1), 1-8.‏

Lee, H. A., Gabriel, R., Bolton, J. P. G., Bale, A. J., & Jackson, M. (2002). Health status and clinical diagnoses of 3000 UK Gulf War veterans. *Journal of the Royal Society of Medicine*, 95(10), 491-497.

Macera, C. A., Aralis, H. J., Highfill-McRoy, R., & Rauh, M. J. (2014). Posttraumatic stress disorder after combat zone deployment among Navy and Marine Corps men and women. *Journal of women's health*, 23(6), 499-505.

Macera, C. A., Aralis, H. J., Highfill-McRoy, R., & Rauh, M. J. (2014). Posttraumatic stress disorder after combat zone deployment among Navy and Marine Corps men and women. *Journal of women's health*, 23(6), 499-505.‏

MacGregor, A. J., Han, P. P., Dougherty, A. L., & Galarneau, M. R. (2012). Effect of dwell time on the mental health of US military personnel with multiple combat tours. *American journal of public health*, 102(S1), S55-S59.

MacGregor, A. J., Tang, J. J., Dougherty, A. L., & Galarneau, M. R. (2013). Deployment-related injury and posttraumatic stress disorder in US military personnel. *Injury*, 44(11), 1458-1464.‏

Maguen, S., Cohen, B., Cohen, G., Madden, E., Bertenthal, D., & Seal, K. (2012). Gender differences in health service utilization among Iraq and Afghanistan veterans with posttraumatic stress disorder. *Journal of Women's Health*, 21(6), 666-673.

Mayo, J. A., MacGregor, A. J., Dougherty, A. L., & Galarneau, M. R. (2013). Role of occupation on new-onset post-traumatic stress disorder and depression among deployed military personnel. *Military medicine*, 178(9), 945-950.

McCarren, M., Janes, G. R., Goldberg, J., Eisen, S. A., True, W. R., & Henderson, W. G. (1995). A twin study of the association of post-traumatic stress disorder and combat exposure with long-term socioeconomic status in Vietnam veterans. *Journal of traumatic stress*, 8, 111-124.‏

Miguel-Tobal, J. J., Cano-Vindel, A., Iruarrizaga, I., Gonza´lez-Ordi, H., & Galea, S. (2005a). Psychopathological repercussions of the March 11 terrorist attacks in Madrid. *Psychology in Spain*, 9, 75–80.

Nandi, A., Galea, S., Ahern, J., & Vlahov, D. (2005). Probable cigarette dependence, PTSD, and depression after an urban disaster: Results from a population survey of New York City residents 4 months after September 11, 2001. *Psychiatry*, 68, 299–310.

Neria, Y., Besser, A., Kiper, D., & Westphal, M. (2010). A longitudinal study of posttraumatic stress disorder, depression, and generalized anxiety disorder in Israeli civilians exposed to war trauma. *Journal of Traumatic Stress, 23*, 322–330.

North, C. S., Nixon, S. J., Shariat, S., Mallonee, S., McMillen, J. C., Spitznagel, E. L., ... Smith, E. M. (1999). Psychiatric disorders among survivors of the Oklahoma City bombing. *Journal of American Medical Association, 282*, 755–762.

North, C. S., Pfefferbaum, B., Kawasaki, A., Lee, S., & Spitznagel, E. L. (2011). Psychosocial adjustment of directly exposed survivors 7 years after the Oklahoma City bombing. *Comprehensive Psychiatry, 52*, 1–8.

North, C. S., Pfefferbaum, B., Narayanan, P., Thielman, S., McCoy, G., Dumont, C., ... Spitznagel, E. L. (2005). Comparison of post-disaster psychiatric disorders after terrorist bombings in Nairobi and Oklahoma City. *British Journal of Psychiatry, 186*, 487–493.

North, C. S., Pfefferbaum, B., Tivis, L., Kawasaki, A., Reddy, C., & Spitznagel, E. L. (2004). The course of posttraumatic stress disorder in a follow-up study of survivors of the Oklahoma City bombing. *Annals of Clinical Psychiatry, 16*, 209–215.

North, C. S., Pollio, D. E., Smith, R. P., King, R. V., Pandya, A., Surı´s, A. M., ... Pfefferbaum, B. (2011). Trauma exposure and posttraumatic stress disorder among employees of New York City companies affected by the September 11, 2001 attacks on the world trade center. *Disaster Medicine and Public Health Preparedness, 5*, 205–213.

Nuttman-Shwartz, O. (2014). Fear, functioning, and coping during exposure to a continuous security threat. Journal of Loss and Trauma, 19(3), 262-277.

Nuttman-Shwartz, O., Dekel, R., & Regev, I. (2015). Continuous exposure to life threats among different age groups in different types of communities. *Psychological Trauma: Theory, Research, Practice, and Policy*, 7(3), 269.

Ohtani, T., Twanami, A., Kasal, K., Yamasue, H., Kato, T., Sasaki, T., ... Kato, N. (2004). Post-traumatic stress disorder symptoms in victims of Tokyo subway attack: A 5-year follow up study. *Psychiatry and Clinical Neurosciences, 58*, 624–629.

O'toole, B. I., Marshall, R. P., Schureck, R. J., & Dobson, M. (1998). Posttraumatic stress disorder and comorbidity in Australian Vietnam veterans: risk factors, chronicity and combat. *Australian and New Zealand Journal of Psychiatry*, *32*(1), 32-42.Palgi, Y. (2015). Predictors of the new criteria for probable PTSD among older adults. *Psychiatry Research, 230*, 777–782.

Palmieri, P. A., Chipman, K. J., Canetti, D., Johnson, R. J., & Hobfoll, S. E. (2010). Prevalence and correlates of sleep problems in adult Israeli Jews exposed to actual or threatened terrorist or rocket attacks. *Journal of Clinical Sleep Medicine, 6*, 557–564.

Paz García-Vera, M., Sanz, J., & Gutierrez, S. (2016). A systematic review of the literature on posttraumatic stress disorder in victims of terrorist attacks. *Psychological reports*, *119*(1), 328-359.‏

Phillips, C. J., LeardMann, C. A., Gumbs, G. R., & Smith, B. (2010). Risk factors for posttraumatic stress disorder among deployed US male marines. *BMC psychiatry*, 10, 1-11.‏

Richardson, L. K., Frueh, B. C., & Acierno, R. (2010). Prevalence estimates of combat-related post-traumatic stress disorder: critical review. *Australian & New Zealand Journal of Psychiatry*, *44*(1), 4-19.‏

Riviere, L. A., Kendall-Robbins, A., McGurk, D., Castro, C. A., & Hoge, C. W. (2011). Coming home may hurt: risk factors for mental ill health in US reservists after deployment in Iraq. *The British Journal of Psychiatry*, 198(2), 136-142.‏

Rona, R. J., Jones, M., Sundin, J., Goodwin, L., Hull, L., Wessely, S., & Fear, N. T. (2012). Predicting persistent posttraumatic stress disorder (PTSD) in UK military personnel who served in Iraq: A longitudinal study. *Journal of psychiatric research*, 46(9), 1191-1198.

Rona, R. J., Jones, M., Sundin, J., Goodwin, L., Hull, L., Wessely, S., & Fear, N. T. (2012). Predicting persistent posttraumatic stress disorder (PTSD) in UK military personnel who served in Iraq: A longitudinal study. *Journal of psychiatric research*, 46(9), 1191-1198.‏

Sandweiss, D. A., Slymen, D. J., LeardMann, C. A., Smith, B., White, M. R., Boyko, E. J., ... & Millennium Cohort Study Team. (2011). Preinjury psychiatric status, injury severity, and postdeployment posttraumatic stress disorder. *Archives of general psychiatry*, 68(5), 496-504.‏

Shalev, A. Y., & Freedman, S. (2005). PTSD following terrorist attacks: A prospective evaluation. *American Journal of Psychiatry, 162*, 1188–1191.

Shear, K. M., Jackson, C. T., Essock, S. M., Donahue, S. A., & Felton, C. J. (2006). Screening for complicated grief among Project Liberty service recipients 18 months after September 11, 2001. *Psychiatric Services*, 57, 1291–1297

Smith, T. C., Ryan, M. A., Wingard, D. L., Slymen, D. J., Sallis, J. F., & Kritz-Silverstein, D. (2008). New onset and persistent symptoms of post-traumatic stress disorder self reported after deployment and combat exposures: prospective population based US military cohort study. *Bmj*, 336(7640), 366-371.

Stein, N. R., Schorr, Y., Krantz, L., Dickstein, B. D., Solomon, Z., Horesh, D., & Litz, B. T. (2013). The differential impact of terrorism on two Israeli communities. *American Journal of Orthopsychiatry*, 83(4), 528-535.

Stuber, J., Resnick, H., & Galea, S. (2006). Gender disparities in posttraumatic stress disorder after mass trauma. *Gender Medicine*, 3, 54–67.

Thompson, W. W., Gottesman, I. I., & Zalewski, C. (2006). Reconciling disparate prevalence rates of PTSD in large samples of US male Vietnam veterans and their controls. *BMC psychiatry*, 6, 1-10.

Toomey, R., Kang, H. K., Karlinsky, J., Baker, D. G., Vasterling, J. J., Alpern, R., ... & Eisen, S. A. (2007). Mental health of US Gulf War veterans 10 years after the war. *The British Journal of Psychiatry*, *190*(5), 385-393.Tracie Shea, M. T., Reddy, M. K., Tyrka, A. R., & Sevin, E. (2013). Risk factors for post-deployment posttraumatic stress disorder in national guard/reserve service members. *Psychiatry research*, 210(3), 1042-1048.

van Liempt, S., van Zuiden, M., Westenberg, H., Super, A., & Vermetten, E. (2013). Impact of impaired sleep on the development of PTSD symptoms in combat veterans: a prospective longitudinal cohort study. *Depression and anxiety*, 30(5), 469-474.‏

Wells, T. S., Ryan, M. A., Jones, K. A., Hooper, T. I., Boyko, E. J., Jacobson, I. G., ... & Gackstetter, G. D. (2012). A comparison of mental health outcomes in persons entering US military service before and after September 11, 2001. *Journal of traumatic stress*, 25(1), 17-24.‏

Xue, C., Ge, Y., Tang, B., Liu, Y., Kang, P., Wang, M., & Zhang, L. (2015). A meta-analysis of risk factors for combat-related PTSD among military personnel and veterans. *PloS one*, *10*(3), e0120270.‏

Zhang, G., North, C. S., Narayanan, P., Kim, Y., Thielman, S., & Pfefferbaum, B. (2013). The course of postdisaster psychiatric disorders in directly exposed civilians after the US Embassy bombing in Nairobi, Kenya: A follow-up study. *Social Psychiatry and Psychiatric Epidemiology, 48*, 195–203.
